# Wearables Anticipate Postoperative Complications: A Prospective Cohort Study

**DOI:** 10.64898/2026.06.02.26354556

**Authors:** Lauren E. Lederer, Ali R. Roghanizad, Thomas Clark Howell, Kimberly Turnage, Dan G. Blazer, Rebecca Knackstedt, E. Shelley Hwang, Jessilyn P. Dunn

**Author notes:** **Corresponding Authors:** E. Shelley Hwang, MD MPH, Seeley Mudd Building 465, DUMC Box 3513, Durham, NC 27710, Jessilyn Dunn, PhD, 534 Research Dr, Durham, North Carolina 27708.

## Abstract

Consumer wearable devices enable continuous passive physiologic monitoring in free-living conditions, yet their capacity to detect early postoperative deterioration following hospital discharge remains poorly characterized. Here we report a prospective observational cohort study evaluating multimodal wearable-derived physiologic signals across the perioperative period in adults undergoing elective oncologic surgery at Duke University Health System.

Participants were monitored using an Oura Ring Gen 2 and Garmin Vivosmart 4 from at least two weeks preoperatively through up to 90 days postoperatively, alongside daily electronic patient-reported pain surveys. Devices captured 3,705 participant-days and 82,833 hours of physiologic data across 46 surgical patients. Oura adherence averaged 21.0 hours/day and was significantly higher than Garmin throughout the study period (17.6 hours/day). Garmin wear time declined significantly following surgery, while Oura adherence remained comparatively stable.

Postoperative complications occurred in 17 participants (37%), including 10 major complications (Clavien–Dindo grade IIIb or higher) with a median onset of 13 days after surgery. Patients with major complications demonstrated significantly greater peak deviations from baseline in the first 10 postoperative days across resting heart rate, sleep temperature deviation, and readiness metrics. In the days before clinically documented major complications, wearable and patient-reported signals diverged from those of participants without major complications, with reduced activity appearing as early as four days before the event, followed by higher reported pain and later elevations in resting heart rate and sleep temperature deviation.

These findings support the feasibility of prolonged perioperative wearable monitoring and suggest that physiologic deterioration preceding major surgical complications may be detectable days before clinical documentation, motivating further development and validation of wearable-based postoperative surveillance strategies.

## Introduction

Over 40 million operative procedures are performed in the US each year, with 15% of surgical patients suffering significant postoperative morbidity, and 1-4% experiencing a perioperative mortality.^1^ Globally, the World Health Organization estimates that perioperative complications contribute to nearly 8 million deaths annually.^1^ At the same time, advances in perioperative care, including enhanced recovery after surgery (ERAS) protocols, have reduced hospital length of stay after major operations. ^2^ While this shift has improved inpatient efficiency, it has also moved more of the recovery period into the home, where physiologic monitoring is limited and postoperative complications may emerge between scheduled clinical encounters. This transition creates a critical monitoring gap after discharge, when patients may be clinically vulnerable but largely unobserved.

A growing body of evidence highlights digital health technologies (DHTs) as a key tool in effective remote patient monitoring (RPM) ecosystems.^3–6^ DHTs can be used to continuously monitor physiological changes, and here, we explore their utility to detect postoperative complications on an outpatient basis following hospital discharge. To date, research has been limited to small feasibility studies. A 2021 systematic review analyzed 45 studies utilizing wearables and digital health technologies (DHTs) for postoperative monitoring.^6^ However, only six studies collected data continuously from patients and only one incorporated a wearable device capable of capturing both activity and cardiovascular metrics.^7^ Most research has focused on narrow endpoints or single sensing modalities, particularly step count and physical activity metrics in orthopedic recovery ^8^, limiting insight into the broader physiologic processes underlying surgical recovery. More recent evidence reflects similar limitations across specialties. Additional studies have demonstrated promise for postoperative complications using wearable devices ^9–11^, yet these efforts remain fragmented and are typically constrained by small sample sizes, short monitoring windows, and limited integration of multimodal physiologic data.

Accordingly, the aim of this study was to evaluate the feasibility and potential clinical utility of continuous perioperative home monitoring using consumer wearable devices. We assessed adherence to wearable device use and daily electronic patient-reported outcome (ePRO) surveys during the perioperative period and examined longitudinal wearable, clinical, and complication data to evaluate physiologic changes in the days preceding major postoperative complications.

## Methods

### Patient Recruitment

This study was approved by the Duke University Health System (DUHS) Institutional Review Board (IRB Protocol #Pro00110892). Adult patients aged 18–75 years with a scheduled colorectal, gastrointestinal, hepatopancreaticobiliary (HPB), or breast reconstruction procedure at Duke University Hospital within 30 days were eligible for participation, with enrollment occurring between 2022-2024. Additional inclusion criteria required access to wireless internet at home and ownership of a smartphone device compatible with the Garmin Connect app and Oura app.

Patients were excluded if they were currently enrolled in another clinical trial involving an interventional treatment or if they did not have access to a smartphone device or reliable internet connectivity.

### Study Design

After providing written informed consent, participants were instructed to download the Oura and Garmin mobile applications to their personal smartphone and were provided with an Oura Ring Gen 2 (Oura Health, Oulu, Finland) and a Garmin Vivosmart 4 wearable device (Garmin Ltd., Olathe, KS).

#### Electronic Patient-Reported Outcomes

(ePROs) consisting of daily pain scores using the Numeric Rating Scale were collected and managed using Research Electronic Data Capture (REDCap) electronic data capture tools hosted at Duke University.^12^ REDCap (Research Electronic Data Capture) is a secure, web-based software platform designed to support data capture for research studies, providing 1) an intuitive interface for validated data capture; 2) audit trails for tracking data manipulation and export procedures; 3) automated export procedures for seamless data downloads to common statistical packages; and 4) procedures for data integration and interoperability with external sources.^13,14^

Participants were asked to wear the wearable devices continuously and complete daily ePROs for a baseline period of at least two weeks prior to surgery and for up to 90 days postoperatively. An early cohort of participants (n=19) were monitored for 30 days postoperatively until the protocol was amended and the rest of the cohort (n=27) were monitored for 90 days postoperatively. Wearable data were collected passively throughout the perioperative period in free-living conditions. No clinical interventions were introduced, and no participants were assigned to experimental groups.

### Data Collection and Storage

Wearable data were collected using the Oura Ring Gen 2, and a Garmin Vivosmart 4 wrist wearable device. The Garmin data were accessed through Fitabase (Small Steps Labs LLC), a secure research platform that enables retrieval of deidentified Garmin wearable data. Oura data were accessed through Oura’s secure research data export platform. Participants were assigned study-specific identifiers to ensure deidentification prior to data access.

Data collected for this study include wearable-derived physiological and behavioral metrics such as daily resting heart rate (RHR) and sleep temperature (Oura), and step count and activity duration (Garmin). Additional parameters include sleep stage distributions (light, deep, REM), total sleep time, sleep efficiency, bedtime and wake time variability, and intraday heart rate and heart rate variability (HRV) profiles. The Oura Ring Gen 2 measures skin temperature using negative temperature coefficient sensors, but only provides a relative measure of an individual’s nightly deviation from their baseline temperature.

Automated data retrieval and preprocessing pipelines were implemented using Python (Python Software Foundation, v3.9+).

Participant demographics and clinical data were extracted from the Duke University Health System electronic health record (EHR). Data collected included age, sex, race and ethnicity, body mass index, smoking history, relevant comorbidities, details of surgical procedure, and perioperative clinical outcomes. Postoperative variables included surgical procedure type, estimated blood loss (EBL), length of initial hospital stay (LOS), total surgical time, postoperative complications, emergency department visits, and hospital readmissions.

### Postoperative Complications

For the purposes of this analysis, a complication was defined as any postoperative EHR-documented clinical complication, including presentation to the emergency department, unplanned hospital readmission, or mortality. Patient-reported symptoms requiring clinical intervention and documented during outpatient postoperative visits or telephone encounters were also classified as complications. The date of each postoperative complication was defined as the date on which symptoms related to the event were first documented in the EHR.

Each complication was independently reviewed and graded according to the Clavien–Dindo classification system based on the type and severity of intervention required.^15^ Events classified as Clavien–Dindo grade IIIb (a complication requiring surgical intervention under general anesthesia) or higher were designated as major complications, while all other complications were classified as minor complications.

### Statistical Analysis

Throughout the analysis, normality was assessed with Shapiro-Wilk tests and Q-Q plots. Where normality is rejected (Shapiro-Wilk p < 0.05) or Q-Q plots show deviation from normality, non-parametric tests were used.

#### Demographics

Participants were classified into three complication groups (Major, Minor, No Complication) from the severity of their highest-grade complication. For Table 1, between-group comparisons were performed as Major versus Minor/No complication combined (two groups: Major vs Other). All tests were two-sided.

**Table 1.** Participant Demographic and Clinical Characteristics Stratified by Postoperative Complication Status. Baseline demographic and perioperative characteristics of the study cohort (N=46) stratified by major complications, minor complications, and no complications. Continuous variables are presented as mean (SD) and median (IQR), and categorical variables are presented as number (percentage). *P* values compare participants who experienced major complications with those who experienced minor or no complications. Surgical procedure types included breast reconstruction, colectomy/proctectomy, and gastric/hepatopancreatobiliary procedures. BMI indicates body mass index; EBL, estimated blood loss; and LOS, length of initial hospital stay.

| Characteristic | Total | Major Complications | Minor | No Complication | P Value (Major vs. Minor/No Complication) |
| --- | --- | --- | --- | --- | --- |
| **Total** | **46** | **10** | **7** | **29** |  |
| **Age** |  |  |  |  |  |
| Age, mean (SD), years | 50.6 (13.6) | 43.9 (16.5) | 53.3 (11.6) | 52.3 (12.6) | 0.07 |
| Age, median (IQR), years | 49.5 (43.0 - 62.0) | 45.0 (32.2 - 47.8) | 52.0 (46.0 - 62.5) | 50.0 (43.0 - 62.0) | 0.07 |
| **Gender** |  |  |  |  | **0.47** |
| Female | 24 (52.2%) | 5 (50.0%) | 6 (85.7%) | 13 (44.8%) |  |
| Male | 8 (17.4%) | 3 (30.0%) | 0 (0.0%) | 5 (17.2%) |  |
| Non-Binary/Gender-Queer/Other | 5 (10.8%) | 1 (10.0%) | 0 (0.0%) | 4 (13.7%) |  |
| Unknown | 9 (19.6%) | 1 (10.0%) | 1 (14.3%) | 7 (24.1%) |  |
| **Sex Assigned at Birth** |  |  |  |  | **0.24** |
| Female | 32 (69.6%) | 5 (50.0%) | 7 (100.0%) | 20 (69.0%) |  |
| Male | 14 (30.4%) | 5 (50.0%) | 0 (0.0%) | 9 (31.0%) |  |
| **Race** |  |  |  |  | **0.7** |
| Asian | 2 (4.3%) | 0 (0.0%) | 1 (14.3%) | 1 (3.4%) |  |
| Black or African American | 6 (13.0%) | 1 (10.0%) | 0 (0.0%) | 5 (17.2%) |  |
| White/Caucasian | 36 (78.3%) | 9 (90.0%) | 5 (71.4%) | 22 (75.9%) |  |
| Unknown | 2 (4.3%) | 0 (0.0%) | 1 (14.3%) | 1 (3.4%) |  |
| **Ethnicity (Hispanic/Latino)** |  |  |  |  | **0.54** |
| Hispanic or Latino | 2 (4.3%) | 0 (0.0%) | 1 (14.3%) | 1 (3.4%) |  |
| Non-Hispanic | 42 (91.3%) | 10 (100.0%) | 5 (71.4%) | 27 (93.1%) |  |
| Unknown | 2 (4.3%) | 0 (0.0%) | 1 (14.3%) | 1 (3.4%) |  |
| **BMI** |  |  |  |  |  |
| BMI, mean (SD), kg/m<sup>2</sup> | 29.2 (6.3) | 29.1 (6.4) | 30.0 (9.5) | 29.0 (5.6) | 0.87 |
| BMI, median (IQR), kg/m<sup>2</sup> | 29.0 (24.0 - 34.0) | 29.0 (24.2 - 34.0) | 30.0 (22.0 - 35.0) | 28.0 (24.0 - 33.0) | 0.87 |
| **Surgical Procedure Type** |  |  |  |  | **0.92** |
| breast reconstruction | 11 (23.9%) | 2 (20.0%) | 1 (14.3%) | 8 (27.6%) |  |
| colectomy/proctectomy | 25 (54.3%) | 6 (60.0%) | 4 (57.1%) | 15 (51.7%) |  |
| gastric/HPB | 10 (21.7%) | 2 (20.0%) | 2 (28.6%) | 6 (20.7%) |  |
| **Total Surgical Time** |  |  |  |  |  |
| Mean (SD), minutes | 372.0 (201.7) | 361.0 (147.1) | 266.0 (146.8) | 401.4 (223.8) | 0.92 |
| Median (IQR), minutes | 335.0 (235.0 - 490.2) | 350.5 (270.8 - 419.5) | 238.0 (177.0 - 341.5) | 352.0 (242.0 - 569.0) | 0.92 |
| **Estimated Blood Loss (EBL)** |  |  |  |  |  |
| Mean (SD), mL | 166.6 (313.3) | 99.0 (71.1) | 64.4 (43.3) | 214.5 (386.2) | 0.92 |
| Median (IQR), mL | 100.0 (31.2 - 150.0) | 100.0 (50.0 - 137.5) | 50.0 (40.0 - 100.0) | 100.0 (30.0 - 200.0) | 0.92 |
| 0-50 mL | 13 (28.3%) | 2 (20.0%) | 2 (28.6%) | 9 (31.0%) |  |
| 50-200 mL | 24 (52.2%) | 7 (70.0%) | 5 (71.4%) | 12 (41.4%) |  |
| 200+ mL | 9 (19.6%) | 1 (10.0%) | 0 (0.0%) | 8 (27.6%) |  |
| **Length of Stay (LOS)** |  |  |  |  |  |
| Mean (SD), days | 3.9 (3.5) | 4.4 (2.0) | 4.3 (4.2) | 3.7 (3.8) | 0.1 |
| Median (IQR), days | 3.0 (2.0 - 4.8) | 4.5 (3.0 - 5.0) | 3.0 (1.5 - 5.0) | 3.0 (2.0 - 4.0) | 0.1 |

Continuous variables (age, BMI, total surgical time, estimated blood loss) were summarized as mean (SD) and median (IQR) by group. Between-group comparisons used the Mann–Whitney U test (Wilcoxon rank-sum test) on the raw continuous values, comparing Major to Minor/No combined. One p-value was produced per variable and was reported once for that variable (e.g. on the mean or median row, or on both summary rows as in your table, depending on journal style). No distributional assumptions were made; the test was used in its standard two-sided form.

Categorical variables (e.g. gender, sex assigned at birth, race/ethnicity, surgical procedure type) were summarized as counts and percentages. Between-group comparisons used the full contingency table (Major vs Other × all category levels). For 2×2 tables, Fisher’s exact test was used; for larger tables, the chi-square test was used. A single p-value per variable was computed and reported on the first row of that variable in the table (e.g. the first category level or the variable label row such as “Race/Ethnicity”); subcategory or additional category rows were left without a p-value to avoid repeating the same test.

#### Adherence Analysis

We aimed to assess wearable device adherence across the study period and stratified by key demographics. Pre-operative and postoperative periods were defined as days < 0 and ≥ 0, respectively (and surgery day (day 0) is excluded where specified). Given that the initial cohort (n=19) completed only 30 days of postoperative monitoring, wear time adherence across the entire cohort was analyzed from study day −10 to 30 with day 0 excluded to accurately assess adherence across both study periods. Within this window, participants contributed data from both devices.

Within-participant comparisons (Garmin vs Oura, or pre-op vs post-op within device) used the Wilcoxon signed-rank test, and between-participant comparisons used the Mann–Whitney U test for two groups (e.g., sex) and the Kruskal–Wallis test for more than two (e.g., day of week, age group). For each participant and device, average wear time was computed across the study duration or per period (pre-op and post-op). Adherence was compared between devices (paired by participant), between pre-op and post-op (paired by participant), by day of week, by sex assigned at birth, and by age group (18–25, 26–41, 42–57, 58–76, 77+ years). One participant’s Oura account was deleted after the study and their relevant adherence and Oura wearable data analysis were removed.

#### Recovery Trajectory Analysis

Recovery was analyzed using daily wearable and clinical data. Participants were classified by whether they experienced at least one major complication (“Major”) or other (only minor complications or no reported complications). Wearable metrics and patient-reported pain were analyzed relative to each participant’s preoperative baseline. For this analysis, the pre-operative baseline period was defined to be between 42 days (6 weeks) before surgery until the day before surgery to define a more stable baseline for participants who had more pre-operative data available. The difference from the individual’s median baseline (value − baseline) was then used to analyze postoperative data for each feature.

Peak burden quantified the maximum postoperative deviation from baseline. For each metric, we calculated the maximum absolute change from baseline (|value − baseline|) occurring within the first 10 postoperative days (postoperative days 0–10). Participants were included if they had at least three postoperative measurement days within this window for the given metric.

If both groups satisfied normality (Shapiro–Wilk P > 0.05), we used Welch’s independent-samples t-test (unequal variances); otherwise we used the Mann–Whitney U test. We tested the null hypothesis of no difference between participants with major complications and those with no or minor complications (Other) using a two-sided significance level of 0.05. Normality of each outcome within groups was assessed using the Shapiro–Wilk test. If both groups satisfied normality (P > 0.05), Welch’s independent-samples t-test was used; otherwise, the Mann–Whitney U test was applied. Results are reported as the test used and the corresponding two-sided P-value.

#### Visualization of Recovery Trajectories

Recovery trajectories were defined by the vector of daily differences from the pre-op baseline (Δ) for each RHR, temperature deviation, steps, sleep duration, deep sleep %, pain, HRV, and readiness. Mean trajectory (Δ mean ± SEM) was plotted for the full cohort and also in a stratified manner (by complication group, EBL, procedure type, surgery time, sex, age [median split]) over two windows: normalized days −10 to 30 and −10 to 90. Trajectories were visually inspected with respect to the baseline period (where Δ = 0), on surgery day (day 0), and at the time of major complications.

#### Complication Analysis

Participants who experienced major complications (Clavien–Dindo grade ≥IIIb occurring on or after postoperative day 6 to ensure enough postoperative data was available to perform the analysis) were aligned by the day of complication, and metric changes were evaluated in the 4 days prior to the event. Participants without major complications were assigned matched postoperative reference days based on the median major complication day across the entire cohort (day 13). Between-group differences in changes from baseline were assessed separately at each time point using Kruskal–Wallis tests.

To visualize longitudinal recovery patterns, group-level daily means and standard errors were calculated for each metric across postoperative days and smoothed using a 3-day rolling mean. Trajectories were displayed for resting heart rate, maximum pain, daily step count, and sleep temperature deviation.

## Results

### Participant Demographics

There were 54 patients enrolled in the study, 46 patients completed surgery and had wearable data available (**Figure 1**; **Table 1**). Across the study period, wearable devices captured 3705 participant-days of data, corresponding to 82,833 hours of physiologic monitoring. Participants were, on average, 50.6 ± 13.6 years of age, and 24 participants (52.2%) identified as female, while thirty-two participants (69.6%) were assigned female sex at birth. Participants were mainly White (36 [78.3%]), with smaller proportions identifying as Black or African American (6 [13.0%]) or Asian (2 [4.3%]); 2 participants (4.3%) identified as Hispanic or Latino ethnicity. Body Mass Index (BMI) was 29.2 ± 6.3.

**Figure 1.**
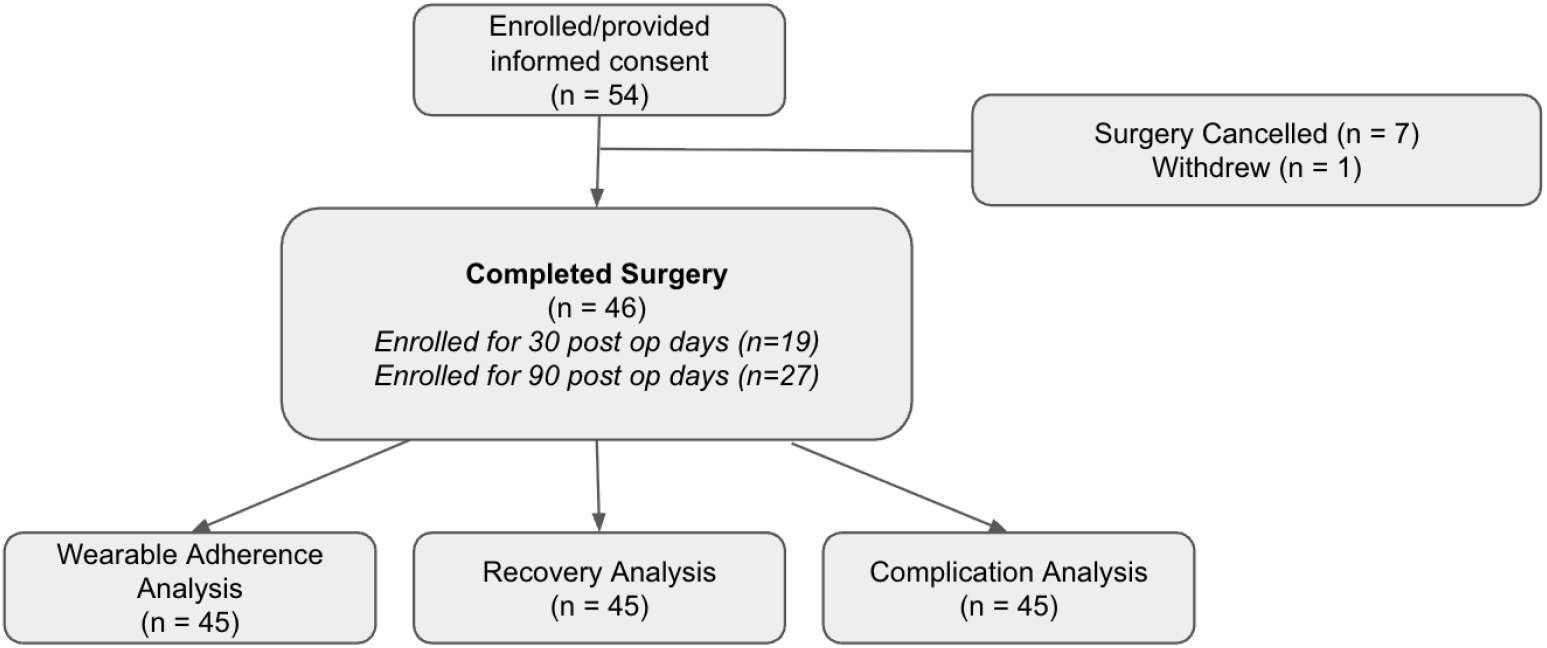
Patient Flow Diagram of study enrollment and data availability. The diagram shows participant flow from initial screening through enrollment, exclusions, follow-up, and inclusion in the final analytic cohort, including exclusions due to incomplete wearable or survey data.

Surgical procedures included colectomy or proctectomy (25 [54.3%]), breast reconstruction (11 [23.9%]), and gastric or hepatopancreatobiliary procedures (10 [21.7%]). Mean operative time was 372.0 ± 201.7 minutes, estimated blood loss was 166.6 ± 313.3 mL, and average LOS was 3.9 ± 3.5 days.

Overall, 10 participants (21.7%) experienced at least one major postoperative complication and 7 (15.2%) other participants experienced only minor complications. Baseline demographic and perioperative characteristics (e.g. surgery type, estimated blood loss, total surgical time) did not significantly differ between participants with major complications and those with minor or no complications (all *P* > 0.05), although participants with major complications tended to be younger (43.9 vs 51.8 years; *P* = 0.07).

### Wearable Device Adherence

Adherence to wearable device use was high (>18 hours/day on average) over the study period spanning 10 days before surgery through 30 days after surgery (**Figure 2A**). Pre-operatively, Garmin was worn 21.59 ± 2.99 hours/day and Oura was worn 21.92 ± 2.53 hours/day. A significant drop in wear time occurred for the Garmin (3.81 hours/day less wear time; Wilcoxon signed-rank test; P < 0.001) (**Figure 2B**). Wear time did not differ significantly by sex, age, race/ethnicity, or day of week (all P > 0.05) (**Figure 2C-D**).

**Figure 2.**
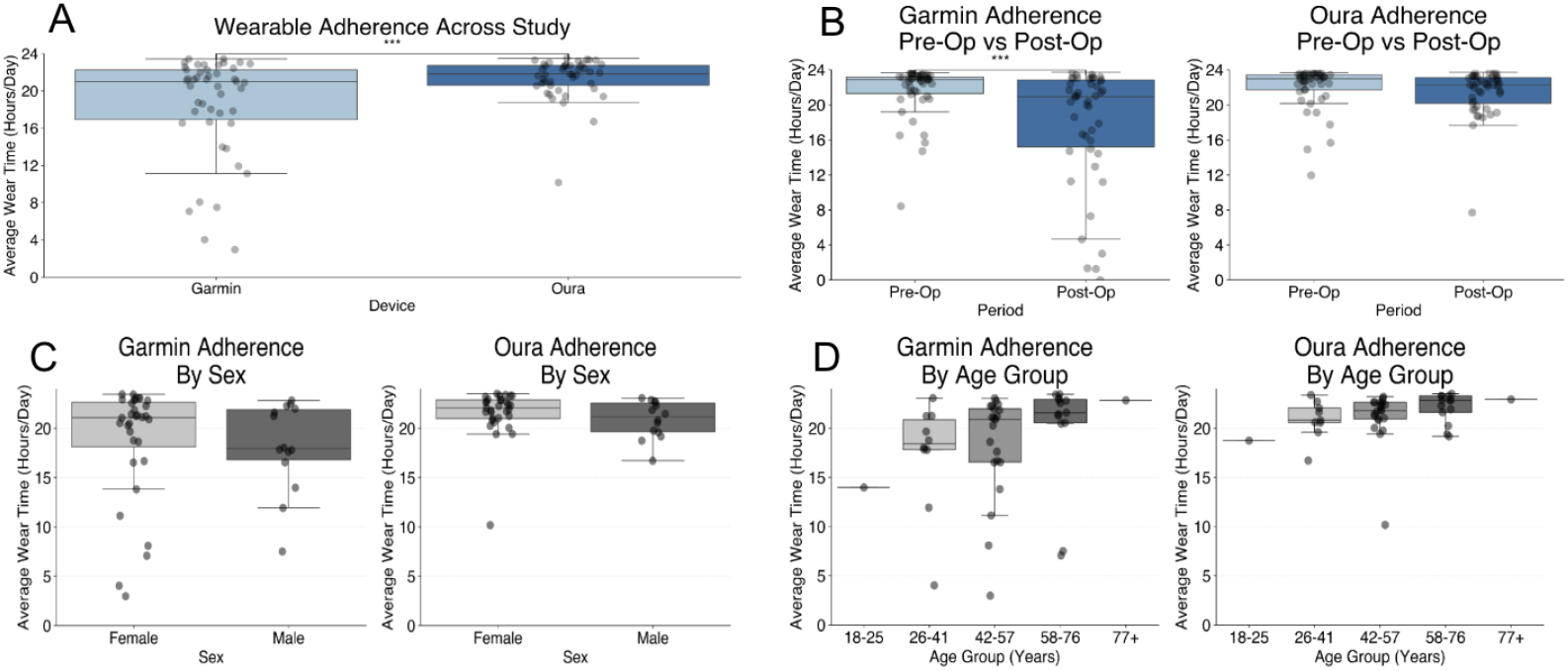
Wearable Device Adherence Comparison. (A) Average daily wear time across the study period for Garmin (n=46) and Oura (n=45 paired participants), demonstrating higher adherence with the Oura device (P<.001). (B) Preoperative versus postoperative average daily wear time by device (excluding the day of surgery), showing a significant postoperative decline in Garmin adherence (n=46 paired participants; P<.001). (C)Average daily wear time stratified by participant sex, with no significant differences observed for either device (P >.05). (D)Average daily wear time stratified by participant age group, with no significant differences observed for either device (P > .05).

### Postoperative Recovery Trajectories

Patients with major complications demonstrated significantly different recovery trajectories from those with minor or no complications across several wearable and survey metrics (**Figure 3**). Trajectory differences are shown for resting heart rate (Oura and Garmin) (**Figure 3A**), maximum daily pain (**Figure 3B**), Oura sleep temperature deviation (**Figure 3C**), steps (Oura and Garmin) (**Figure 3D**). In the first 10 postoperative days, peak absolute deviation from baseline was higher in the major-complication group for Garmin resting heart rate (22.2 ± 7.1 vs 14.5 ± 7.1 bpm; P=0.005, Mann–Whitney U), Oura resting heart rate (26.2 ± 12.5 vs 17.7 ± 10.1 bpm; P=0.046, Mann–Whitney U), Oura sleep temperature deviation (1.53 ± 0.81 vs 1.00 ± 0.54 °C; P=0.043, Mann–Whitney U), and Oura readiness score (43.4 ± 15.6 vs 29.7 ± 12.4; P=0.034, Welch t-test).

**Figure 3.**
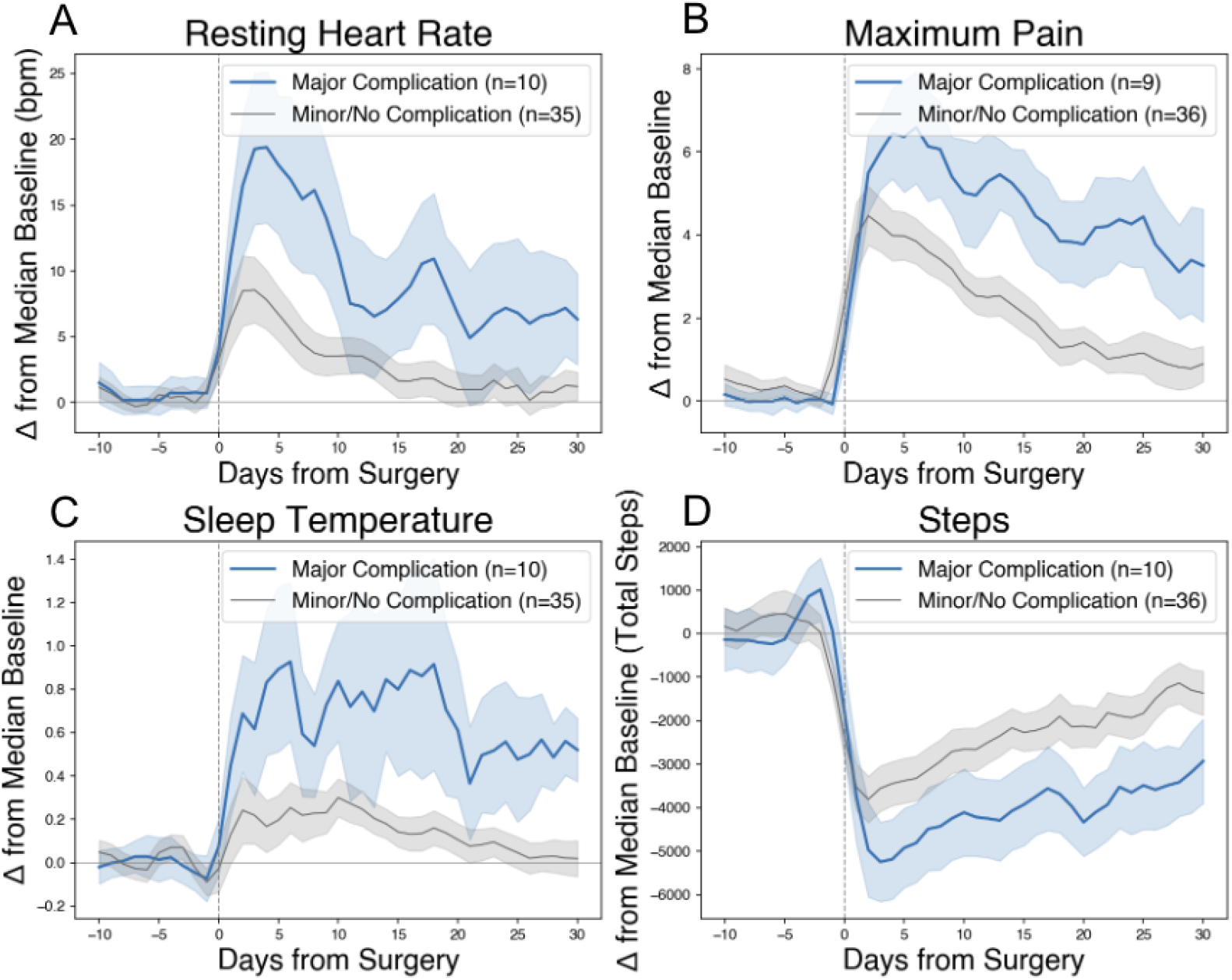
Recovery Trajectories for Patients with Major Complications. Mean trajectories (solid lines) and 95% confidence intervals (shaded areas) are shown for patients with major complications (blue) and minor or no complications (gray) across postoperative days, with values expressed as change from preoperative baseline. (A) Resting heart rate, (B) maximum pain, (C) sleep temperature, and (D) daily step count are displayed. Vertical dashed lines indicate the day of surgery (postoperative day 0). Across all metrics, patients with major complications exhibited slower and more prolonged deviations from baseline compared with patients with minor or no complications. For each postoperative day, group-level means and standard errors of the mean (SEM) were computed across all participants with available preoperative baseline values and valid daily measurements for the corresponding metric. To improve visualization of longitudinal trends, daily mean trajectories and SEM were smoothed within each complication group using a 3-point centered rolling mean (window = 3 days; minimum observations = 1).

### Changes in Wearable and Clinical Metrics Preceding Major Complications

Participants who experienced major postoperative complications showed greater physiologic and behavioral deviations from preoperative baseline in the days preceding the event than participants with minor or no complications (**Figure 4**). Across the four days before the complication (days 4, 3, 2, and 1), group differences emerged in a time-dependent pattern. Daily steps were significantly lower among participants with major complications beginning four days before the event and persisting through day 2 (day 4: −6766 vs −3117 steps, P=0.021; day 3: −5560 vs −3212 steps, P=0.049; day 2: −5730 vs −3066 steps, P=0.008) (**Figure 4C**). Maximum pain scores were significantly higher in the major complication group at days 3 and 2 before the event (7.0 vs 2.0 at both time points; day 3: P=0.04; day 2: P=0.006) (**Figure 4B**). One day before the complication, resting heart rate and sleep temperature deviation were both significantly elevated in the major complication group compared with the minor/no complication group (resting heart rate: 13.2 vs 4.2 bpm, P=0.04; sleep temperature deviation: 0.85 vs 0.17°C, P=0.02) (**Figure 4A and 4D**).

**Figure 4.**
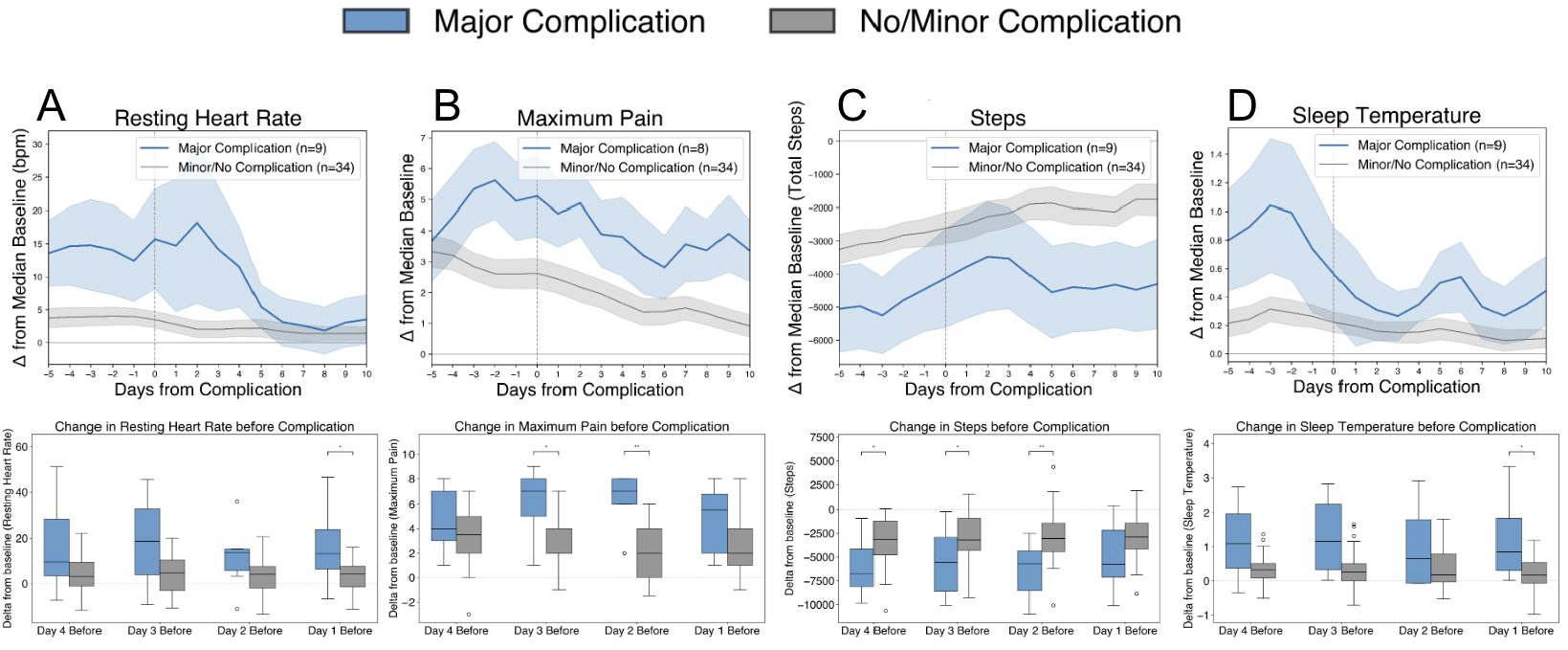
Change in wearable metrics preceding major postoperative complications. Boxplots show changes from preoperative baseline (median of days −10 to −1) for (A) resting heart rate, (B) maximum pain score, (C) daily step count, and (D) sleep temperature deviation at 4, 3, 2, and 1 days before complication events. Participants with major complications (Clavien–Dindo ≥IIIb; blue) were compared with those with minor or no complications (gray) using matched postoperative reference days. Line plots show postoperative trajectories expressed as change from preoperative baseline for participants with major complications (blue) and minor or no complications (gray). Vertical dashed lines indicate the day of surgery (postoperative day 0).

Additional significant differences were observed. Oura readiness score was significantly lower in the major complication group at day 4 (−33 vs −12 points, P=0.04) and day 1 (−32 vs −9 points, P=0.03). Sleep duration was significantly greater in the major complication group at day 3 (1.1 vs −0.03 hours relative to baseline, P=0.03). Resting heart rate differences at days 4, 3, and 2 before the event did not reach statistical significance (P>0.05 for all comparisons).

## Discussion

This prospective cohort study demonstrates both the feasibility and potential clinical relevance of continuous perioperative monitoring using consumer wearable devices in free-living surgical patients. Across 46 surgical participants, wearable devices captured 3,705 participant-days and 82,833 hours of physiologic monitoring, with high adherence sustained throughout home recovery. Major postoperative complications occurred in 10 participants (21.7%) and were associated with more severe and prolonged physiologic recovery disturbances compared with participants experiencing minor or no complications. Measurable deviations in wearable data, including resting heart rate, increased sleep temperature deviation, and reduced steps, emerged in the days preceding clinically documented major complications.

These findings extend a growing body of literature supporting digital health technologies for postoperative remote patient monitoring. Prior systematic reviews and meta-analyses have concluded that wearable monitoring in surgical populations is feasible but remains limited by heterogeneous methodologies, short monitoring periods, and reliance on narrow endpoints such as step count or activity alone. ^3,4,8^ Much of the existing literature has focused on orthopedic recovery or short-duration activity monitoring, providing limited insight into the broader physiologic processes underlying postoperative recovery and deterioration. Our findings support these prior observations while contributing longitudinal multimodal physiologic data collected continuously under free-living conditions across multiple surgical specialties.

The observed physiologic patterns preceding complications are biologically plausible and consistent with the known surgical stress response.^16,17^ Surgery induces a coordinated neuroendocrine and inflammatory response characterized by sympathetic activation, altered thermoregulation, metabolic stress, and reduced physical activity. Elevated resting heart rate may reflect increased physiologic stress, autonomic imbalance, pain, or early systemic inflammation, while increased sleep temperature deviation may reflect inflammatory or infectious processes that precede overt clinical presentation.^18^ Reduced activity and persistent pain may similarly represent impaired recovery or early symptom emergence.^19,20^

Device adherence findings provide additional insight relevant to real-world deployment of wearable-based monitoring systems. Both devices achieved high adherence overall (>17 hours on average across the study period), although Oura wear time remained significantly higher than Garmin throughout the perioperative period. The ring-based form factor, nocturnal wear characteristics, or differences in user experience and data presentation may partially explain this observation. While the present study was not designed to determine the cause of adherence differences, these findings suggest that device selection and user experience may substantially influence long-term monitoring success and should be considered in future implementation studies.

Taken together, these findings support the concept that postoperative recovery and deterioration are continuously measurable using wearable-derived physiologic data. Rather than replacing existing postoperative care pathways, wearable monitoring may provide a complementary layer of physiologic surveillance during the transition from hospital to home. Future work should focus on validating multimodal digital biomarkers and determining whether interpretable wearable-derived signals can improve early detection, risk stratification, and clinical decision-making during postoperative recovery.

## Limitations

Several limitations should be considered when interpreting these findings. First, this was a single-center observational cohort study with a limited number of major complications. Although sufficient to characterize feasibility and identify physiologic associations, the study was not powered to validate predictive models or establish clinically actionable thresholds.

Second, wearable data completeness varied across participants and time periods. Some participants intermittently discontinued device wear, particularly in the postoperative period, introducing potential bias and reducing analyzable participant-days. Because behavioral changes such as device removal may occur concurrently with physiologic deterioration, distinguishing biologic signal from monitoring behavior remains an ongoing challenge and represents an important consideration for future studies.^21^

Third, the timing of complications was determined from electronic health record documentation rather than physiologic onset. Symptoms and physiologic deterioration may begin several days before formal clinical documentation, introducing temporal uncertainty when aligning wearable changes to complication events. Future studies incorporating real-time symptom reporting and clinician adjudication may improve temporal precision.

Fourth, the wearable devices used in this study provide proprietary, device-derived metrics rather than raw physiologic signals, and certain measurements reflect individualized baseline changes rather than absolute physiologic values. Consequently, observed physiologic changes should be interpreted as wearable-derived correlates of recovery rather than direct clinical measurements.

Finally, the study cohort consisted of patients undergoing elective oncologic and reconstructive procedures at a single academic medical center. Generalizability to emergency procedures, non-oncologic surgery, or broader healthcare settings remains uncertain and warrants investigation in larger multicenter cohorts.

## Conclusions

Continuous perioperative monitoring using consumer wearable devices was feasible across a diverse surgical cohort and generated high-resolution physiologic data throughout home recovery. Patients who developed major postoperative complications demonstrated prolonged physiologic recovery disturbances compared with those experiencing minor or no complications. Importantly, wearable-derived physiologic and behavioral deviations, including deviations from baseline resting heart rate, increased sleep temperature, increased sleep duration, reduced activity, and increased pain, were significantly different in the days preceding clinically documented major complications.

Together, these findings suggest that postoperative recovery is continuously measurable using wearable-derived physiologic data and support further investigation of wearable-based postoperative surveillance strategies. Future work should focus on validating multimodal digital biomarkers and developing interpretable risk prediction models capable of identifying early physiologic deterioration and supporting timely clinical intervention during home recovery.

## Acknowledgments

We thank Kaileigh Moertl, Stephanie Sullivan, Stacy Murray, Adi Molvin, and Melissa Rodgman for their valuable contributions to the conduct of this study.

## Ethics Approval and Consent to Participate

This study was approved by the Duke University Health System Institutional Review Board (IRB Protocol #Pro00110892). All participants provided written informed consent.

## Data Availability

Data may be made available from the corresponding author upon reasonable request and subject to applicable institutional approvals, data use agreements, and participant privacy protections.

## References

1. Dobson, G. P. Trauma of major surgery: A global problem that is not going away. Int. J. Surg. Lond. Engl. 81, 47–54 (2020).

2. Melnyk, M., Casey, R. G., Black, P. & Koupparis, A. J. Enhanced recovery after surgery (ERAS) protocols: Time to change practice? Can. Urol. Assoc. J. 5, 342–348 (2011).

3. Khanna, A. K., Flick, M. & Saugel, B. Continuous vital sign monitoring of patients recovering from surgery on general wards: a narrative review. Br. J. Anaesth. 134, 501–509 (2025).

4. Tan, S. Y., Sumner, J., Wang, Y. & Wenjun Yip, A. A systematic review of the impacts of remote patient monitoring (RPM) interventions on safety, adherence, quality-of-life and cost-related outcomes. Npj Digit. Med. 7, 192 (2024).

5. Pandit, J. A., Pawelek, J. B., Leff, B. & Topol, E. J. The hospital at home in the USA: current status and future prospects. NPJ Digit. Med. 7, 48 (2024).

6. Dawes, A. J., Lin, A. Y., Varghese, C., Russell, M. M. & Lin, A. Y. Mobile health technology for remote home monitoring after surgery: a meta-analysis. Br. J. Surg. 108, 1304–1314 (2021).

7. Breteler, M. J. M. et al. Wireless Remote Home Monitoring of Vital Signs in Patients Discharged Early After Esophagectomy: Observational Feasibility Study. JMIR Perioper. Med. 3, e21705 (2020).

8. Hall III, R. R., Hamel, A. P., Chan, P. A., Zheng, H. & Ayers, D. C. Use of Wearable Devices to Augment Traditional Measurements of Postoperative Outcomes Following Total Joint Arthroplasty: Systematic Review. JMIR Rehabil. Assist. Technol. 13, e84671 (2026).

9. Hua, R. et al. Biorhythms derived from consumer wearables predict postoperative complications in children. Sci. Adv. 11, eadv2643 (2025).

10. Beqari, J. et al. A Pilot Study Using Machine-learning Algorithms and Wearable Technology for the Early Detection of Postoperative Complications After Cardiothoracic Surgery. Ann. Surg. 281, 514 (2025).

11. Zhang, J. et al. Predicting Post-Operative Complications with Wearables: A Case Study with Patients Undergoing Pancreatic Surgery. Proc. ACM Interact. Mob. Wearable Ubiquitous Technol. 6, 87:1-87:27 (2022).

12. Williamson, A. & Hoggart, B. Pain: a review of three commonly used pain rating scales. https://onlinelibrary.wiley.com/doi/10.1111/j.1365-2702.2005.01121.x.

13. Harris, P. A. et al. The REDCap consortium: Building an international community of software platform partners. J. Biomed. Inform. 95, 103208 (2019).

14. Harris, P. A. et al. Research electronic data capture (REDCap)--a metadata-driven methodology and workflow process for providing translational research informatics support. J. Biomed. Inform. 42, 377–381 (2009).

15. Clavien, P. A. et al. The Clavien-Dindo classification of surgical complications: five-year experience. Ann. Surg. 250, 187–196 (2009).

16. Cusack, B. & Buggy, D. J. Anaesthesia, analgesia, and the surgical stress response. BJA Educ. 20, 321–328 (2020).

17. Giannoudis, P. V., Dinopoulos, H., Chalidis, B. & Hall, G. M. Surgical stress response. Injury 37, S3–S9 (2006).

18. Pan, W.-T., Ji, M., Ma, D. & Yang, J.-J. Effect of perioperative autonomic nervous system imbalance on surgical outcomes: a systematic review. Br. J. Anaesth. 135, 608–622 (2025).

19. Nevo, Y. et al. Activity Tracking After Surgery: Does It Correlate With Postoperative Complications? Am. Surg. 88, 226–232 (2022).

20. Hanley, C., Ladha, K. S., Clarke, H. A., Cuthbertson, B. C. & Wijeysundera, D. N. Association of postoperative complications with persistent post-surgical pain: a multicentre prospective cohort study. Br. J. Anaesth. 128, 311–320 (2022).

21. Lederer, L. et al. The Importance of Data Quality Control in Using Fitbit Device Data From the All of Us Research Program. JMIR MHealth UHealth 11, e45103 (2023).

